# Cannabis potential effects to prevent or attenuate SARS-COV2 contagion

**DOI:** 10.1101/2022.03.31.22273181

**Authors:** M. Herrera-Gómez Paula, F. Echeverri-Cataño Luis, S Giraldo, Y Ruiz Colorado-, Alberto Vélez van Meerbeke

## Abstract

Medical cannabis has gained an exponential interest in recent years. Therapeutic targets have been broadened from specific applications over pain control, chemotherapy side effects, treatment-resistant epilepsies and multiple sclerosis, among others. Several in vitro and animal studies, along with few human controlled studies, suggest cannabinoids have a potential therapeutic role over medical conditions comporting inflammatory mechanisms. Given the tremendous world-wide impact of the COVID-19 pandemic, research efforts are converging towards the use of cannabinoids to attenuate severe or fatal forms of the disease. The present survey aims to explore possible correlations between cannabis use, either recreational or medical, over the presence of SARS-COV-2 contagion, along with the symptom’s severity. 4026 surveys were collected via electronic form. Results suggest a relation between any type of cannabis use and a lower risk of SARS-COV-2 contagion (*p=*0,004; OR=0,689, IC95% 0,534-0,889). Despite several methodological limitations, the present survey steps up the urge to expand our understanding on cannabinoids potential use on human controlled studies, that can better arm us in the fight against the current COVID-19 pandemic.

## Introduction

Cannabinoids have been regarded as a possible therapeutic clue for the SARS-COV-2 pandemic. Even though, till date there is limited evidence towards a specific therapeutic role for cannabinoids on viral infections ^1^. Cannabinoids anti-inflammatory mechanisms have been only explored during recent years, mainly tested in vitro, in vivo, animals and computational settings^2,3^. Comparatively, human controlled studies are still scarce^4^.

THC and CBD anti-inflammatory effects are highly suspected given their therapeutic potential use in diverse conditions having a physio pathological core around inflammation and immune cells^1,5–7^. Based on available literature, it is reasonable to think that cannabinoids may play multiple therapeutic roles in front of SARS-COV2 contagion, as well as on other viral infections. These perspectives embrace a potential preventive role through replication inhibition mechanisms, and a direct therapeutic role linked to a possible modulation of angiotensin converting enzyme 2 (ACE2) ^8–10^ and cytokines cascades ^11^, both related to the most severe and fatal SARS-COV-2 issues ^3^.

The present survey aims to explore possible correlations between cannabis use, either recreational or medical, over the presence of SARS-COV-2 contagion, along with the symptom’s severity.

## Methods

### Study design and population

We carried out a cross-sectional study through a self-administered online survey among a non-probabilistic sample in Colombia. The survey remained online from 1^st^ to 30^th^ of November, 2020.

### Sample size calculation and sampling strategy

We calculated sample size with an expected frequency of 50%, an acceptable error of 5%, and a design effect of 1 for a population survey, of a minimum sample size of 384 participants. Sampling was obtained with a virtual snowball method, by posting the survey in social media groups and contacts (WhatsApp©, Telegram®, Facebook®) asking to answer and share the link with multiple contacts.

We did not use any imputation method after trying to collect the information. After giving their agreement, participants were allowed to answer the survey: *1. Age, 2. “Have you been diagnosed with COVID-19?”, 3. “If you have had COVID-19, what was the intensity of your symptoms?”, 4. “Has any of the people you live with, or with whom you have very close contact, been infected with COVID-19?”, 5. “Please indicate the number of people close to you who have been infected”, 5*.*”Do you use or consume any type of cannabis?”, 6. “If you answered ‘yes’, please indicate which type (recreational, sativa oil, indica oil, hemp mother tincture, topic or uncontrolled preparations)”, 7. “How frequently do you use these products? (daily, occasionally)”*.

After completing the scheduled data collection, a visual inspection was conducted to verify the database completion. A semi-automated procedure was applied to homogenize outputs. Statistical analyses were conducted on IBM SPSS Statistics for Mac, Version 26.0. Data distribution was checked to verify sphericity, skewness and kurtosis. Parametric analysis (univariate and bivariate analysis) through Chi-square test and risk estimation were conducted.

### Ethical considerations

This research is supported by the Ethics Committee from Universidad Tecnologica de Pereira (Comité de Bioética de la Universidad Tecnologica de Pereira), who gave the ethical approval (# 52-28062). Once participants entered the survey link, they were explicitly told that participation is voluntary and data gathered will be processed anonymously and solely for investigative purposes. Participants were not asked to give their name or any contact information.

## Results

4026 surveys were collected through an online questionnaire. Three participants did not fill the age information. Data obtained was retained to apply statistical analysis. The mean age was 34.9 years (SD 11.6; min 18, max 86). 2272 participants declared any type of cannabis use (56,4%); 727 (18,4%) declared confirmed SARS-COV-2 contagion by test or by highly suggestive symptoms. Among those participants under the category *SARS-CoV2 positive or suspected infection*, 10,3% were cannabis users, and 7,8% were non consumers. Referring to the type of medicinal cannabis employed, participants mainly declared the use of uncontrolled preparations like topical ointments (67,9%), followed by Cannabis sativa oil (43,9%) and hemp mother tincture (12,4%). 341 participants declared recreational and medical use jointly (See Table 1 for full data).

**Table 1.** Cannabis consumption profile.

|  | Total | Recreational use |  | Medical use |  |
| --- | --- | --- | --- | --- | --- |
|  |  | n | % | n | % |
| **CANNABIS USE DISTRIBUTION** | 2272 | 1822 | 100% | 784 | 100% |
| Recreational and medical use jointly |  | 341 | 18,7% | 341 | 85,0% |
| Mother tincture |  | 37 | 2,0% | 97 | 12,4% |
| Mother tincture and oils |  | 0 | 0,0% | 8 | 1,0% |
| Hemp (*indica*) oil |  | 0 | 0,0% | 6 | 0,8% |
| Sativa *sativa* oil |  | 195 | 10,7% | 344 | 43,9% |
| Topical ointments |  | 231 | 12,7% | 532 | 67,9% |
| One or more | Yes | 1481 | 81,3% | 359 | 45,8% |
|  | No | 341 | 18,7% | 425 | 54,2% |
| Frequency of use | Daily | 1160 | 63,7% | 335 | 42,8% |
|  | occasional | 661 | 36,3% | 447 | 57,2% |

In regard to the question *“Do you have any member of your family or people with whom you live under the same roof, that has been positive to SARS-COV2 contagion?*”, almost 60% of the participants expressed at least one contagion among close cohabitants and 40% expressed living with more than 1 person with a confirmed infection (mean number of contagions = 1.8 *(sd = 0*.*7)*, range between 1 to 150).

62% of participants declared having had low or mild intensity symptoms, 6% required hospitalization and 0,83% required ICU management. (See Figure 1 and 2 for full data description).

**Figure 1.** Symptoms severity among cannabis users Vs non users.

**Figure 2.** SARS-COV-2 contagion frequencies according to cannabis consumption in a Colombian sample, 2020.

Bivariate analysis was conducted, examining all variables in study (Cannabis consumption, confirmed infection, presence of symptoms, severity of symptoms, close-related people with SARS-COV2 confirmed or suspected infection, type of cannabis if consumed, recreational or medical use of cannabis, simultaneous use, quantity and frequency of cannabis if used). Patients who use cannabis have a lower risk of SARS-COV2 infection with an OR of 0.689 (95% CI 0.534-0.889) and this association is statistically significant (*p*=0.004). No adjustments were made in the OR since only one of the Chi-square tests yielded significant results.

## Discussion

The present study found that there is a significant statistical association between any type of cannabis use and a lower risk of SARS-COV-2 contagion (*p*=0,004; OR=0,689, IC95% 0,534-0,889). No other relations nor associations were found in the population survey.

Despite the estimated sample size declared in Methodology (N=384), total data obtained from online surveys was used to complete the statistical analysis (N=4026). Given the conditions declared about the non-probabilistic sampling, the association found in the statistical analysis only can be suggested, not assured, due to selection bias. Another limitation is the absence of data in regard to doses quantification and biomarkers of endocannabinoid response in individuals consuming cannabis having SARS-CoV2 positive or suspected infection, in order to compare effects among different populations, dosing quantification, doses-response relation, biological and sociodemographic characteristics.

These considerations could be taken into account for future research under more rigorous experimental protocols.

Despite our survey limitations, we can consider a reasonable amount of evidence pointing at the phytocannabinoids possible mechanisms to prevent contagion or attenuate symptoms severity.

The SARS-CoV-2 initial contagion enters the host through spike proteins (SP) having a high affinity with angiotensin-converting-enzyme 2 (ACE2) and transmembrane protease serine 2 (TMPRSS2). Both proteins are expressed in multiple tissues, especially in lung, urogenital and gastrointestinal tissue, and participate in the fusion of the virus envelope with the the target cells membranes through an endocytosis mechanism. Inside the cell, the mRNA virus is released, completing the transcription and translation process of the viral structures through the ribosome, packaged in the Golgi apparatus, and infecting the contiguous cells by exocytosis ^12^.

Cytokine release syndrome is triggered by internalization of ACE2 and activation of angiotensin 2, which in turns activates the expression of KB nuclear factor, increasing proinflammatory cytokines (IL-6, TNFα, IL-1β, and IL-10). Among the main described effects, there is a blood pressure elevation and impaired lung function ^13^.

Furthermore, as demonstrated in macrophages and on cells culture from lung epithelia, SARS-CoV-2 SP can induce a proinflammatory response in infected cells by triggering inflammasome NLRP3 (NOD-like receptor Pyrin domain containing 3) complex activation. NLRP3 protein, as part of pattern recognition receptors (PRR), is a NOD-like (nucleotide-binding oligomerization domain) receptor containing the pyrin effector domain able to trigger and participate in inflammasome formation. During SARS-CoV-2 infection, this pathway is hyperactivated, leading to an aberrant release of cytokines and other proinflammatory molecules at the root of cellular pyroptosis, and consequent cytokine storm that features poor prognosis in COVID-19 patients. Considering the well-established “gate-keeper” properties of cannabidiol (CBD) due to its ability to rescue RhoA GTPase, it has been proposed as a possible molecule having the capacity to downregulate NLRP3/Caspase-1/IL-1β and IL-18 pathway ^14^.

It has been suggested that high-cannabidiol *sativa* extracts are able to down-regulate the expression of the two key receptors for SARS-CoV-2 in several models of human epithelia ^17^. The CB2 receptor agonists, is the main therapeutic target of cannabinoids, including immunomodulation effects, blocking virus entry and viral replication inhibition. When CB2 receptor is activated, it occurs specific proteins downward regulation (ACE2 and TMPRSS), thus preventing the SARS-CoV-2 invasion, which in turn, blocks viral replication, inhibits the migration of macrophages, and promotes the change of the proinflammatory phenotype of macrophages (M2) triggered by infection, towards anti-inflammatory phenotype characterized by release of IL10 and TGF-b.

Virus replication can be prevented by the malfunction of different Ca2+-dependent enzyme complexes implicated in inflammatory processes, including transglutaminases, calpains, and matrix metalloproteinases. The CB1 receptor agonists induced by Anandamide (AEA) and 2-arachidonoylglycerol (2-AG), can inhibit Ca2+ ions release, stabilizing function of Ca2+-dependent proteins and changing signal transduction ^18.^

In spite of the promising studies describing anti-inflammatory and antioxidative properties ^15, 16^, there is still a long way ahead to fully grasp the phytocannabinoids pharmacological properties. Particular attention must be paid in regard to potential adverse effects, depending on specific phytocannabinoids, the type of extract or compounds, and the administration pathway. For instance, even if their incidence is low, CBD-mediated impairment of immune response may increase the risk of pneumonia, viral infection or boosting of an already existing one ^19^. Other considerations should be taken with those patients having hepatic enzymes alterations, like those exposed to antiepileptic or retroviral medications ^20^. Further research is always needed, even though available reports on medical cannabis keep on throwing lights on their enormous therapeutic potentials, with a particular hope as a powerful tool to cope with our global health challenge in front of the COVID-19 pandemic and its lasting post-infections effects.

## Data Availability

All data produced in the present study are available upon reasonable request to the authors

## Notes

### Competing Interest Statement

The authors have declared no competing interest.

### Funding Statement

This study did not receive any funding

### Author Declarations

This research is supported by the Ethics Committee from Universidad Tecnologica de Pereira (Comite de Bioetica de la Universidad Tecnologica de Pereira), who gave the ethical approval (# 52-28062)

